# Influenza Vaccine Effectiveness Against Pediatric Deaths: 2016–2025

**DOI:** 10.64898/2026.02.20.26346732

**Authors:** Jerome S. Leonard, Katie Reinhart, Peng-Jun Lu, Tammy A. Santibanez, Anup Srivastav, Mei-Chuan Hung, Anurag Jain, Alicia Budd, Stacy Huang, Krista Kniss, Ashley M. Price, Erin Burns, Sascha Ellington, Brendan Flannery

## Abstract

**BACKGROUND AND OBJECTIVES:** Seasonal influenza vaccination has been shown to reduce the risk of influenza and severe complications among children 6 months and older. Since 2010, reported numbers of influenza-associated pediatric deaths among children aged <18 years have ranged from 37 during the 2011–2012 season to 289 during 2024–2025. We estimated influenza vaccine effectiveness (VE) against pediatric death from 2016–2017 through 2024–2025.

**METHODS:** We conducted a case-cohort analysis comparing current season influenza vaccination status among reported influenza-associated pediatric deaths with survey estimates of influenza vaccination coverage in pediatric age groups. Underlying medical conditions and current seasonal influenza vaccination were obtained from surveillance case reports. We estimated vaccination odds ratios (OR) and 95% confidence intervals (CI) from logistic regression comparing influenza vaccination among children who died with vaccination coverage in comparison cohorts. VE was calculated as (1 – OR) × 100.

**RESULTS:** From August 2016 through July 2025, 1234 laboratory-confirmed influenza-associated pediatric deaths were reported among children aged 6 months--17 years. Of 1086 reported deaths including influenza vaccination information, 124 (23%) of 530 children with underlying medical conditions and 70 (13%) of 556 children without known conditions were fully vaccinated against influenza. Average influenza vaccination coverage in survey cohorts was 49%. VE was 80% (95% CI, 75% to 84%) overall, 77% (95% CI, 71% to 82%) among children with underlying medical conditions and 87% (95% CI, 84% to 89%) among children without known conditions.

**CONCLUSIONS:** Influenza vaccination reduced risk of fatal influenza among children with or without known underlying medical conditions.

## INTRODUCTION

Seasonal influenza vaccination has been recommended to reduce the risk of influenza and its severe complications, including influenza-associated death in children and adolescents.^1,2^ While rare, influenza-associated pediatric deaths occur annually with varying incidence depending upon the influenza season.^3,4^ Children younger than 5 years and children and adolescents with certain underlying medical conditions are at increased risk of hospitalization and complications attributable to influenza.^5^ Neurologic, cardiac, pulmonary conditions and genetic disorders have been associated with higher risk for influenza-associated pediatric death.^3,6,7^ However, children with no underlying conditions still constitute approximately half of influenza-associated pediatric deaths.^4,6-9^

In the United States, deaths in children and adolescents with laboratory-confirmed influenza have been reportable since 2004.^3^ From 2005 through 2009, seasonal influenza was estimated to account for more than 100 deaths annually among children and adolescents.^10-12^ During the 2009 influenza A(H1N1) pandemic, 288 influenza-associated deaths in children and adolescents were reported.^3,13^ Since 2010, reported numbers of influenza-associated deaths among children aged <18 years have ranged from 37 in the 2011–2012 season to 289 in the 2024–2025 season (as of January 2026). The number of influenza-associated pediatric deaths reported during the 2024–2025 season was the highest since reporting started.^3,4^ Since 2021, US influenza vaccination rates have been in decline, leaving more children susceptible to severe complications or death due to influenza.^14^ In this report, we evaluated influenza vaccine effectiveness against influenza-associated pediatric deaths among children with and without known underlying medical conditions over eight influenza seasons.

## METHODS

### Data Source

We analyzed deaths reported to the US Influenza-Associated Pediatric Mortality Surveillance System during 4 seasons before the start of the COVID-19 pandemic (2016–2017 through 2019–2020), and four seasons from 2021–2022 through 2024–2025. During the 2020–2021 season, influenza activity was reduced and only one influenza-associated pediatric death was reported to the Centers for Disease Control and Prevention (CDC). For this analysis, influenza seasons began on epidemiologic week 40 and ended on week 39 of the following year (approximately October through September). This system has been described in detail previously.^6-8,15-17^ Briefly, influenza-associated deaths are defined as deaths resulting from a clinically compatible illness in US residents aged <18 years with laboratory-confirmed influenza virus infection, with no period of complete recovery between the illness and death. Confirmation of influenza virus infection included clinical diagnostic testing during illness or post-mortem examination using antigen detection, viral culture or nucleic acid amplification.

State and local health departments reported influenza-associated pediatric deaths to CDC using a standard case report form transmitted over secure, Web-based interface. Case reports include sex, birthdate (or age), underlying medical conditions, current season influenza vaccination status and date of most recent influenza vaccination (if known), state of residence at time of illness, date of illness onset, date of hospital admission (if hospitalized), clinical information and complications during fatal illness, laboratory tests performed and results (including influenza testing as well as tests for viral and bacterial co-infection), location of death (emergency department, hospital, during transportation to hospital, or out of hospital), and date of death.

State and local health department personnel documented underlying medical conditions from medical records and coroners’ reports. Underlying medical conditions associated with increased risk for influenza-related complications among children and adolescents included asthma, chronic lung disease (including cystic fibrosis), neurologic and neurodevelopment conditions, blood disorders (including sickle cell disease), endocrine disorders (including diabetes mellitus), heart disease (including congenital heart disease), kidney disorders, liver disorders, metabolic disorders (including mitochondrial disorders), obesity (sex-specific body mass index [BMI] above the 95^th^ percentile for ages 2-18 years), pregnancy, long-term therapy with aspirin- or salicylate-containing medications, weakened immune systems due to diseases (including HIV or AIDS, leukemia and other cancers) or immunosuppressive medications (including chemotherapy and chronic corticosteroids), and disabilities involving muscle function, lung function, or difficulty coughing, swallowing, or clearing fluids from airways.^5^ For VE analyses, we stratified pediatric deaths into two groups: children with ≥1 underlying medical condition associated with increased risk of complications from influenza and children with no known high-risk condition. Deaths among children with incomplete medical histories or with health conditions not specified as higher risk of influenza complications^5^ (history of febrile seizures, skin and soft tissue infections, and other conditions) were grouped with deaths among children with no known underlying medical condition.

To classify current and prior influenza vaccination status, health department personnel determined vaccination status based upon review of available information from state or county immunization information systems, case patient medical records, health care providers, parental or coroner reports. Children were considered fully vaccinated if they had received one or two doses of current season influenza vaccine as recommended by the Advisory Committee on Immunization Practices^1^ at least 14 days before onset of illness. Children aged 6 months through 8 years were considered partially vaccinated if they were previously unvaccinated and received one dose of current season influenza vaccine ≥14 days before onset of fatal illness or a second dose <14 days before illness onset. For case reports without illness onset date, we estimated onset as five days before death based on the median duration of illness among cases with recorded date of symptom onset. Children who were partially vaccinated were included as unvaccinated for demographic purposes (Table 1) but excluded from vaccine effectiveness calculations (Figure 2 and 3). Children who had been vaccinated in prior seasons without current season vaccination were considered unvaccinated for the season of fatal infection. In primary analyses, we excluded cases in which health department personnel could not determine the child’s current season vaccination status.

**TABLE 1.**
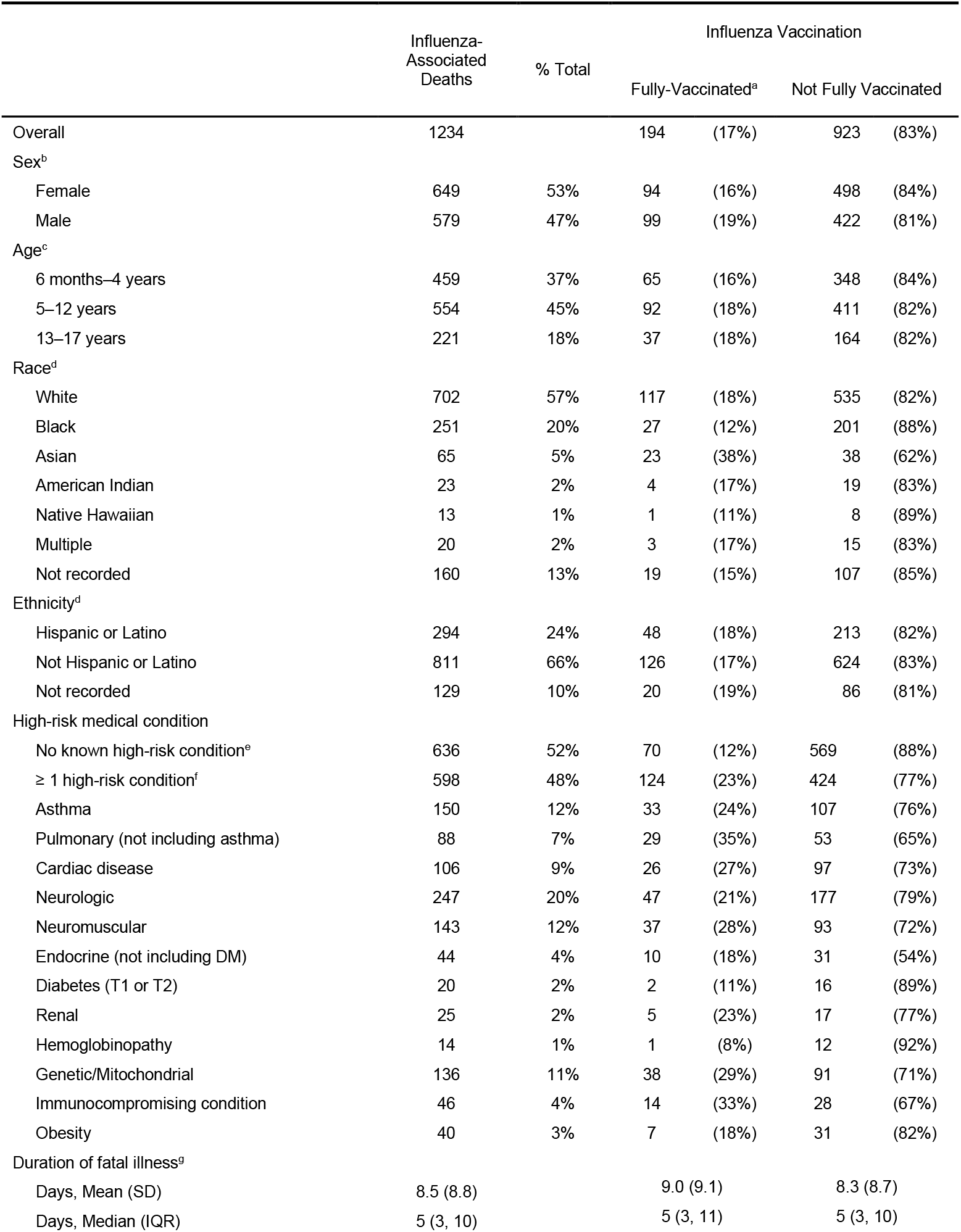
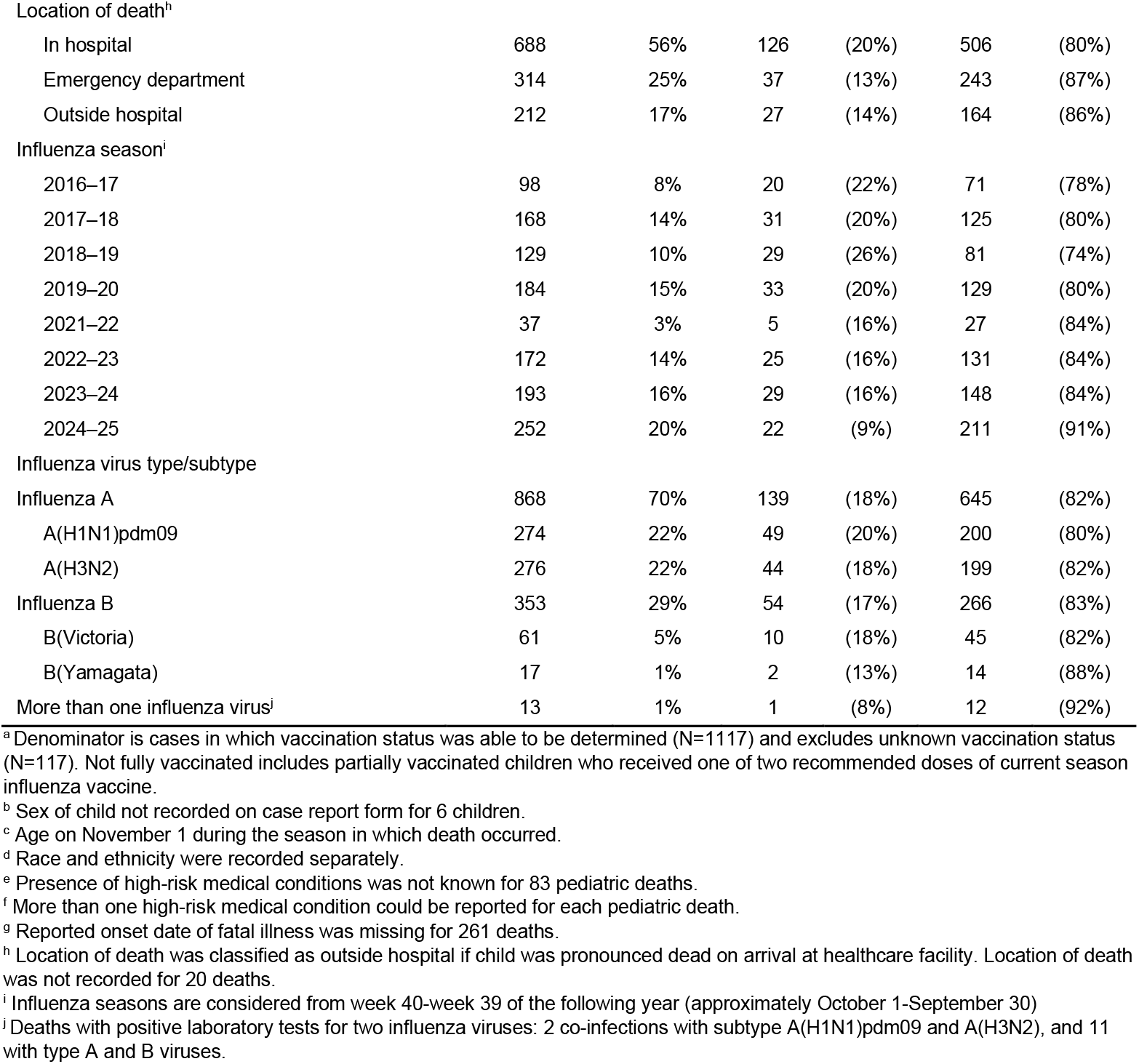
Pediatric influenza-associated deaths and percent with documented influenza vaccination, October 2016-September 2025.

For consistency with influenza vaccination coverage estimates, age groups were defined based on the child’s age on November 1 of the influenza season when death occurred. Because influenza vaccines are not approved for use in children aged <6 months, reported deaths among children aged <6 months on November 1 were excluded.

### Influenza Vaccination Coverage in Cohorts

We used two sources of influenza vaccination coverage among comparison cohorts of U.S. children and adolescents: National Immunization Survey-Flu (NIS-Flu),^18^ and National Health Interview Survey (NHIS).^19^ NIS-Flu is a national telephone survey of households with children aged 6 months through 17 years that produces national and state-level estimates (and for some cities, including Chicago and New York City) of influenza vaccination coverage among children. NHIS is an in-person household survey that provides nationally representative estimates of influenza vaccination for children aged 6 months through 17 years. Data from NIS-Flu and NHIS are weighted by age, sex, race/ethnicity and geographic area to represent the US population. Cumulative estimates of seasonal influenza vaccination coverage are estimated from parental report of child’s receipt of ≥1 dose of seasonal influenza vaccine since July 1. Monthly coverage estimates and standard errors for vaccination received during July or August (depending upon survey and season) through May were calculated by Kaplan-Meier survival analysis of data from interviews conducted from August, September or October through June.^20-22^ Age group was based upon the child’s age on November 1 of each influenza season. NHIS was used to estimate ratios of vaccination coverage among children in the survey sample identified as having specified health conditions or without these conditions. From 2016–2018, NHIS instruments asked whether the child or adolescent had experienced an asthma episode in the past 12 months, ever been diagnosed with diabetes mellitus, cystic fibrosis, sickle cell anemia, congenital heart disease or other heart condition, cerebral palsy, muscular dystrophy or seizures. From 2018–2025, NHIS instruments asked only whether the child or adolescent had experienced an asthma episode in the past 12 months or ever been diagnosed with diabetes mellitus. Comparison of influenza vaccination coverage during 2016–2018 among children in the NHIS sample with any specified health conditions versus asthma or diabetes mellitus yielded similar coverage ratios.

We defined comparison cohorts for each influenza-associated death by age category (6 months–4 years, 5–12 and 13–17 years) and state of residence. We obtained estimated mean and standard deviation of influenza vaccination coverage in the calendar month preceding case illness onset for each influenza season from publicly available NIS-Flu data.^6^ To account for differences in vaccination coverage among US children with underlying medical conditions compared with survey averages, we multiplied NIS-Flu comparison coverage estimates for cases with high-risk conditions by ratio of cumulative influenza vaccination coverage among children with, versus without underlying medical conditions from the annual NHIS survey by age category and season. Due to changes in collection of high-risk conditions in 2019 and availability of NHIS data through 2024, average ratios for 2016–2020 and from 2023–2024 were applied to NIS-Flu comparison coverage estimates for the 2018–2019 and 2024–2025 seasons, respectively.

### Influenza Vaccine Effectiveness

To estimate influenza vaccine effectiveness against influenza-associated death, we adapted the screening method based on a case-cohort analysis as previously described.^2,23,24^ The case-cohort method produces an odds ratio (OR), which estimates the relative risk for influenza-associated death in fully-vaccinated children versus those who were not vaccinated against influenza. We included children at least 6 months old on November 1 of the influenza season during which death occurred who were eligible to receive at least one dose of seasonal influenza vaccine. We calculated ORs and 95% confidence limits comparing odds of current season influenza vaccination among cases with odds of vaccination in comparison cohorts by age category, state of residence and calendar month preceding case illness onset. We used logistic regression with case vaccination status as the dependent variable and the log odds of current season influenza vaccination (proportion vaccinated / 1 – proportion vaccinated) in the NIS-Flu comparison cohort entered as an offset. With this offset in the model, the intercept provides an estimate of the log(OR). Influenza VE was estimated as (1 – OR) x 100, expressed as a percent. For children with high-risk conditions, likelihood of vaccination was based on NHIS-adjusted seasonal influenza vaccination coverage among children aged 6 months to 18 years with underlying medical conditions, by age group, season and calendar month.

In sensitivity analyses, we evaluated the impact on VE of assuming that cases with undetermined influenza vaccination status were either all vaccinated or all unvaccinated, and evaluated the impact on VE of decreasing vaccination coverage in comparison cohorts by 10 and 20 percent to account for over-reporting of child and adolescent influenza vaccination in national survey data.^25-28^ Analyses were conducted in R (version 4.5.1; R Core Team, Vienna, Austria).

This activity was reviewed by CDC, deemed not research, and was conducted in accordance with applicable federal law and CDC policy.

## RESULTS

### Influenza-Associated Pediatric Deaths

From October 2016 through September 2025, excluding the 2020–2021 season, 1234 influenza-associated pediatric deaths among children aged 6 months to 17 years were reported by 48 states, Chicago, New York City, and Washington D.C. The number of pediatric deaths reported per season among children aged over 6 months ranged from 37 during 2021–2022 to 252 during 2024–2025 (Table 1). Overall, 598 (48%) pediatric deaths occurred in children with ≥1 underlying medical conditions and 636 (52%) in those without known conditions. More of the influenza-associated deaths in older children and adolescents had underlying medical conditions (51% among those aged 5–12 and 56% among 13–17 years) compared with 36% among deaths in children aged 6 months–4 years. Location of death was reported for 1214 cases: 688 (56%) died during hospitalization, 314 (25%) in the emergency department, and 212 (17%) out of hospital. In all, 868 deaths were associated with influenza A (274 H1N1pdm09, 276 H3N2, and 318 with unknown subtype), 353 deaths were associated with influenza B, and 13 deaths had infection with more than one virus type or influenza A subtype. Mean and median number of days from illness onset to death was 9.0 and 5 days, respectively, among vaccinated children and 8.3 and 5 days among unvaccinated children.

### Influenza Vaccination among Pediatric Deaths

Among 1117 influenza-associated deaths in children with influenza vaccination information, the percent of children who were fully vaccinated against influenza ranged from 16% to 19% by age group, sex and ethnicity, with wider range by race (38%) (Table 1). By influenza season, the percent of children with current season influenza vaccination ranged from 26% during 2018–2019 to 9% during 2024–2025 (Figure 1). Across all seasons, 18% of fatal influenza A and 17% of influenza B-associated cases occurred among children who were fully vaccinated against influenza, with similar percentages by influenza A virus subtype. Among deaths in children without known conditions, 12% were fully vaccinated against influenza compared with 23% among deaths in children with ≥1 underlying medical conditions. Among deaths in children with ≥1 high-risk medical conditions, the percent that had been vaccinated cases was highest among those with non-asthma pulmonary disorders (35%), followed by immunocompromising conditions (33%) and genetic and mitochondrial disorders (29%). The lowest percentages of influenza-associated deaths in children who had been fully vaccinated against influenza were among those with hemoglobinopathies (8%), diabetes (11%) and obesity (18%).

**FIGURE 1.**
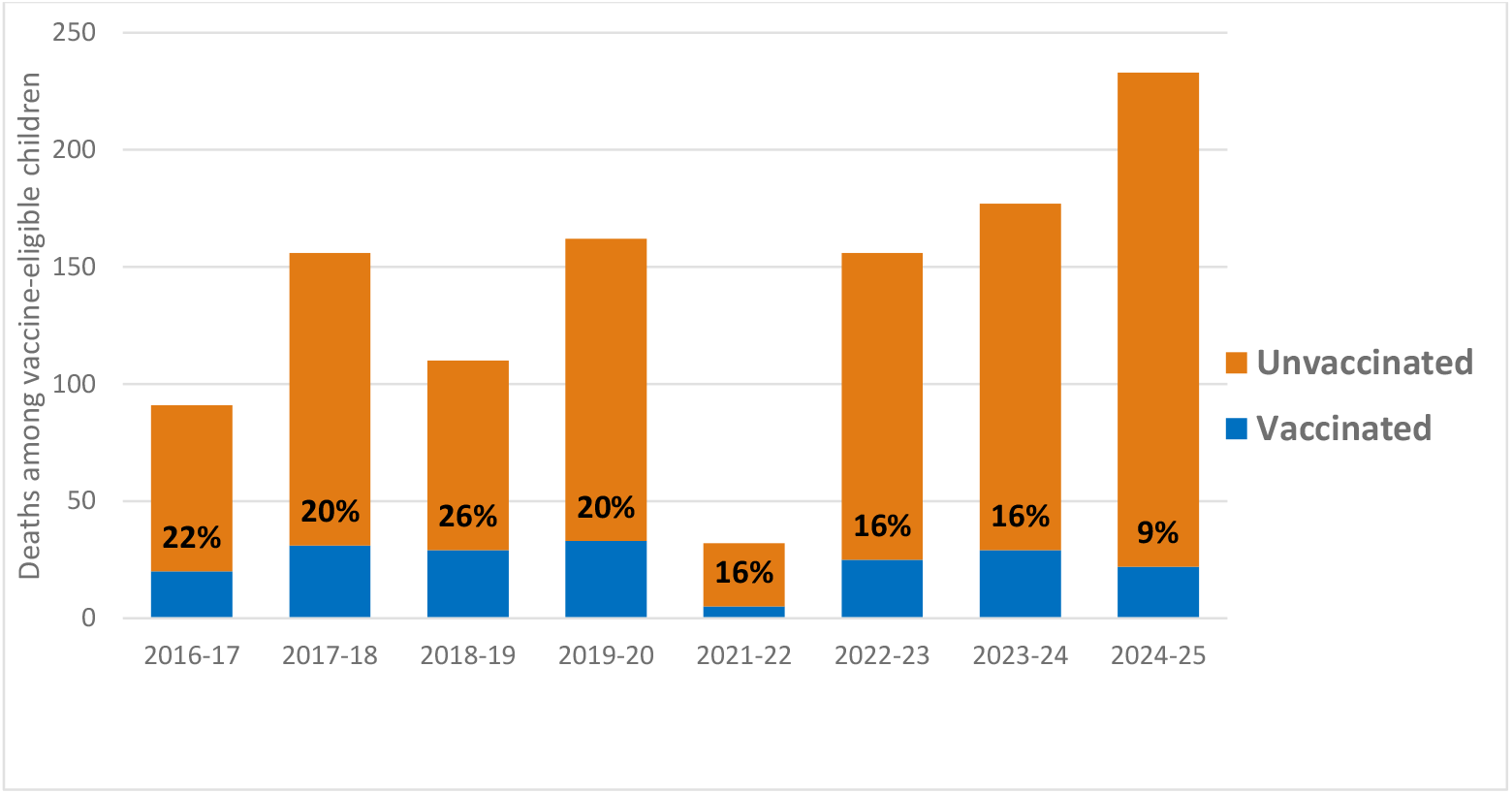
Number and Percent of Influenza-Associated Pediatric Deaths in Influenza Vaccinated or Unvaccinated US Children and Adolescents Aged 6 Months to 17 Years, 2016–2017 Through 2024–2025 (Excluding 2020–2021) Influenza Seasons. Numbers of deaths are shown among children with or without documented or reported influenza vaccination for the season in which death occurred, and percent of deaths among children fully vaccinated against influenza. Data were reported to the Pediatric Mortality Surveillance System.

### Influenza Vaccination Coverage Estimates

During 2016–2020, mean survey estimates of current season influenza vaccination coverage in comparison cohorts ranged from 50% to 58% among children with specified underlying medical conditions, versus 51% to 59% among children without known conditions. During 2021–2025 after the first waves of COVID-19, mean survey estimates in comparison cohorts decreased, ranging from 42% to 50% among children with high-risk medical conditions, versus 39% to 48% among children without known conditions.

### Influenza Vaccine Effectiveness

After excluding deaths among children who were partially vaccinated, 194 (18%) of 1086 influenza-associated deaths occurred in children with current season influenza vaccination and 892 (82%) in unvaccinated children. Overall, VE against influenza-associated pediatric death was 80% (95% CI: 75-84) (Figure 2). VE against death was higher among children without known high-risk conditions (87%; 95% CI: 84-89) than among children with high-risk medical conditions (77%; 95% CI: 71-82). Among children with high-risk medical conditions, VE estimates were higher for those aged 6 months–4 years (83%; 95% CI: 78-88) than those aged 13–17 years (66%; 95% CI: 57-73). VE against influenza A-associated death was 68% (95% CI: 61-74) compared with 77% (95% CI: 72-81) against influenza B-associated death, with overlapping confidence intervals. Among children without known underlying medical conditions, VE estimates were also higher for those aged 6 months–4 years (89%; 95% CI: 86-91) compared with those aged 13–17 years (80%; 95% CI: 74-84). VE was 87% (95% CI: 84-90) against pediatric deaths associated with influenza A and 87% (95% CI: 84-89) against deaths associated with influenza B. Among children with high-risk medical conditions, influenza VE against pediatric deaths each season ranged from 66% (95% CI: 58-72) in 2019–2020 to 84% (95% CI: 81-87) in 2024–2025. Among children without known high-risk conditions, VE each season ranged from 77% (95% CI: 72-81) in 2022–2023 to 95% (95% CI: 93-96) in 2024–2025.

**FIGURE 2.**
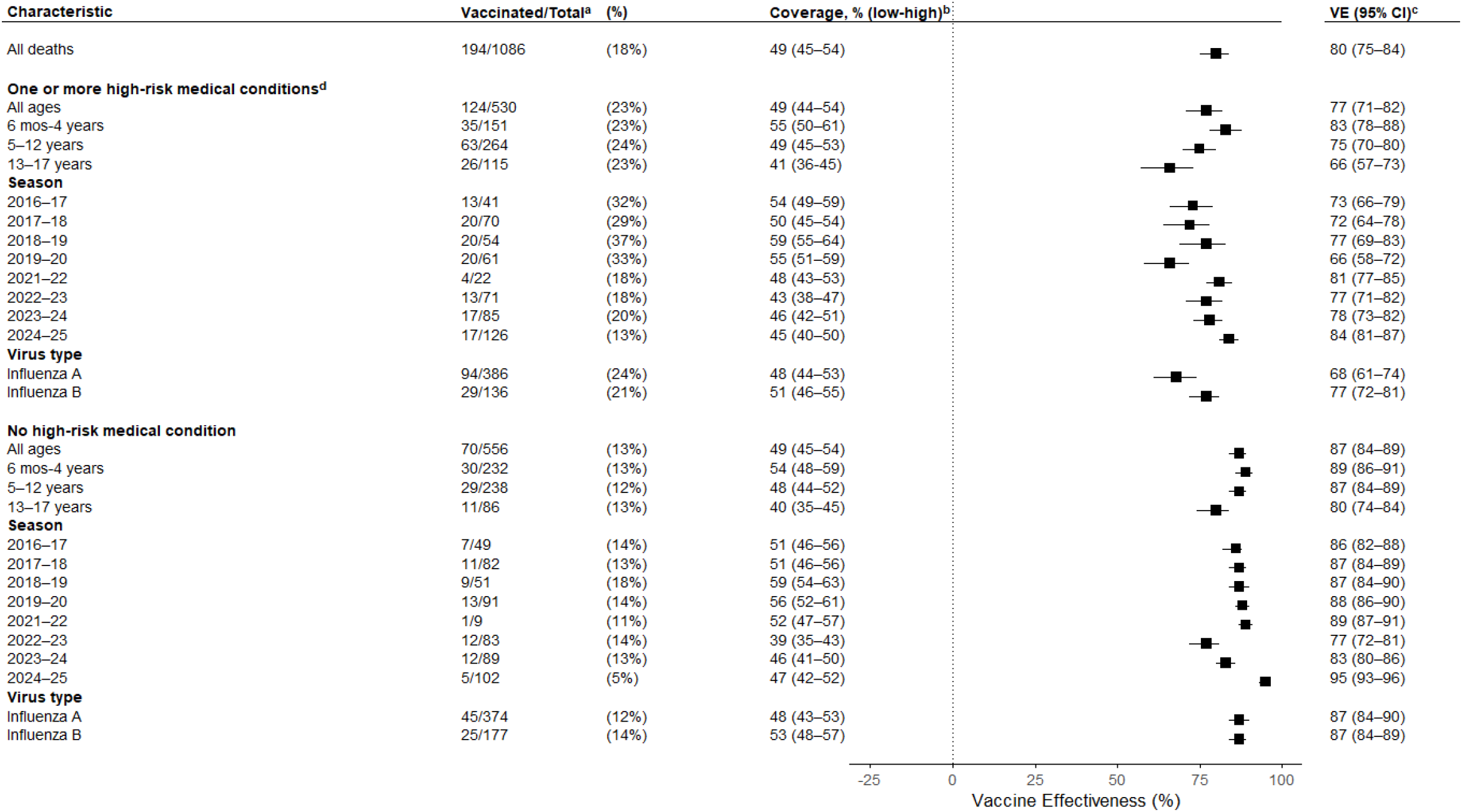
Influenza Vaccine Effectiveness Estimates and Percentage Vaccinated Among Influenza-Associated Pediatric Deaths Compared With NIS-Flu Survey Influenza Vaccination Coverage Estimate, by Season and Age Group VE, vaccine effectiveness; CI, confidence interval. ^a^ Excludes 31 cases with partial vaccination (receipt of one of two recommended doses in the season during which death occurred) or documented vaccination <14 days before onset of fatal illness, and 117 cases with vaccination status listed as unknown on the case report form. ^b^ Average influenza vaccination coverage among children in National Immunization Survey-Flu (NIS-Flu) sample by case age category, state, season and reference month. For deaths among children with at least one documented high-risk medical condition, NIS-Flu coverage was multiplied by the ratio of influenza vaccination coverage in the National Health Interview Survey (NHIS) among children with specified high-risk conditions compared with children in the survey sample without high-risk conditions by age category, state influenza vaccination coverage category (low, medium or high) and season (see Methods). ^c^ Percent vaccine effectiveness = 100 x (1 - Vaccination Odds Ratio), estimated as odds of vaccination (p / (1-p)) among cases divided by odds of vaccination (coverage / (1-coverage)) in comparison group, with 95% confidence intervals. ^d^ Documentation of at least one high-risk medical condition on case report form. Case report forms for 83 pediatric deaths indicated that underlying medical conditions were unknown.

Sensitivity analyses assuming that all cases with undetermined vaccination status were fully vaccinated resulted in lower but statistically significant estimates of 66% (95% CI: 58-73) among children with high-risk medical conditions and 74% (95% CI: 68-79) among children without known high-risk conditions (Figure 3). Assuming that cases with unknown vaccination status were not vaccinated had minimal impact on VE estimates. When seasonal influenza vaccination coverage estimates among comparison cohorts were decreased by 10 and 20 percent, VE against death among children with high-risk conditions was 63% (95% CI: 56-69) and 54% (95% CI: 46-61), respectively. Among children without known high-risk conditions, decreasing comparison coverage estimates by 10 or 20 percent, VE against death was 84% (95% CI: 80-86) and 79% (95% CI: 76-82). VE estimates by age category remained statistically significant in all sensitivity analyses.

**FIGURE 3.**
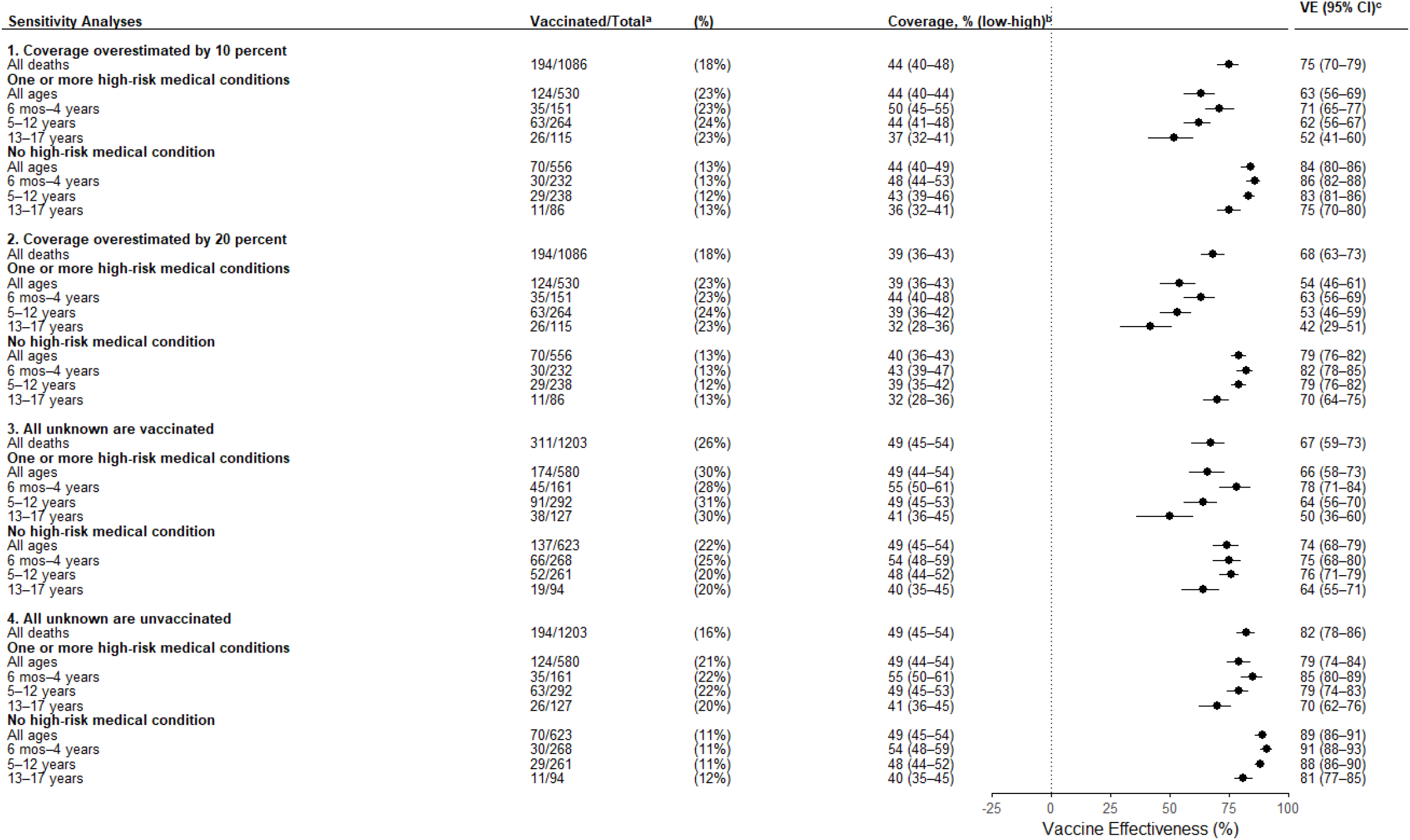
Sensitivity Analyses for Over-Estimation of Influenza Vaccination Coverage in NIS-Flu Survey and Unknown Vaccination Status Among Pediatric Deaths. VE, vaccine effectiveness; CI, confidence interval. ^a^ Number of deaths among fully vaccinated children (excluding deaths among partially vaccinated children) varies by sensitivity analysis: (1) and (2) exclude deaths among children with vaccination status listed as unknown; (3) re-classified all deaths among those with unknown vaccination status as fully vaccinated; (4) re-classified all deaths among those with unknown vaccination status as unvaccinated. ^b^ In sensitivity analysis (1), comparison coverage for every influenza-associated death by case age category, state, season and reference month was decreased by 10%, assuming 10% parental over-report in National Immunization Survey-Flu (NIS-Flu) sample, and in analysis (2) comparison coverage for each death was decreased by 20%, assuming 20% parental over-report in NIS-Flu. For deaths among children with at least one documented high-risk medical condition, comparison coverage estimates were decreased by 10% or 20% after NIS-Flu coverage was multiplied by the National Health Interview Survey (NHIS) influenza coverage ratio among children with or without specified high-risk conditions by age category, state influenza vaccination coverage category (low, medium or high) and season. ^c^ Percent VE and 95% CI were estimated separately for each sensitivity analysis as VE = 100 x (1 - Vaccination Odds Ratio), estimated as odds of vaccination (p / (1-p)) among cases divided by odds of vaccination (coverage / (1-coverage)) in comparison group, after applying assumptions to deaths with unknown vaccination status or decreasing comparison coverage estimates to account for potential over-reporting in NIS-Flu.

## DISCUSSION

In this analysis, pediatric influenza-associated deaths were less likely to have been fully vaccinated against influenza than US children in the same age group and state of residence. Of pediatric deaths in children aged ≥6 months, fewer than one in five were fully vaccinated against influenza, and fewer than one in four among deaths in children and adolescents with underlying medical conditions associated with increased risk of severe influenza and complications. Approximately half of the pediatric deaths occurred in children without known health conditions, only 12% of whom were fully vaccinated against influenza. In comparison groups of US children and adolescents, influenza vaccination coverage was consistently higher than among children who died with influenza-associated complications. In sensitivity analyses accounting for potential overestimation of influenza vaccination coverage in US survey data, influenza vaccination remained strongly associated with protection. Taken together, results provide additional evidence that seasonal influenza vaccination reduces the risk of influenza-associated deaths in all pediatric age groups.

In addition to protection against influenza-associated deaths,^2^ influenza vaccination has been shown to provide protection against laboratory-confirmed influenza-associated hospitalizations and severe outcomes. A meta-analysis of observational case-control studies that used patients with negative molecular influenza test results for comparison reported pooled VE of 67% against influenza-associated hospitalizations among infants and children.^29^ In multiple US studies conducted from 2016–2017 through 2024–2025, influenza vaccination has been associated with prevention of severe outcomes,^30^ life-threatening influenza virus infection,^31^ influenza-associated hospitalizations, emergency department, urgent care and healthcare visits among children with and without high-risk medical conditions.^32-37^ Previous analysis of pediatric deaths during four seasons from 2010–2014 showed low influenza vaccination rates among children and adolescents who died with influenza compared with state and age-specific vaccination coverage.^2^ In the current analysis of pediatric deaths over eight influenza seasons, influenza vaccination rates among influenza-associated pediatric deaths were consistently low despite prevalence of underlying conditions associated with high risk of severe influenza and complications.^2,4,6-8^

Sensitivity analyses supported our main findings. In the first sensitivity analyses, we decreased comparison cohort vaccination coverage by 10 and 20 percent to account for over-reporting of children’s influenza vaccination by parents and caregivers in NIS-Flu surveys.^38^ In a US pediatric hospitalization network, validation of parental report of their child’s seasonal influenza vaccination overestimated vaccination rates by 18-25%,^27^ which is similar to estimates from national immunization surveys in Australia.^28^ The resulting lower VE estimates remained statistically significant for all age categories among pediatric deaths in children with and without high-risk medical conditions. In the second sensitivity analysis, we included deaths in children for whom vaccination status could not be determined, assuming that all or none had been fully vaccinated. State and local health department personnel searched multiple sources of influenza vaccine information to identify any record or report of vaccination by providers, coroners, parents and caregivers. Assuming that none of the cases with undetermined vaccination status had been fully vaccinated resulted in similar VE estimates. Alternatively, assuming that all were fully vaccinated resulted in lower but still statistically significant estimates. A strength of the current analysis was the ability of state and local health department personnel to link death records to immunization information systems. Since publication in 2017 of influenza vaccine effectiveness against influenza-associated pediatric deaths in the United States,^2^ completeness of vaccine data in the Influenza Associated Pediatric Mortality Surveillance System has improved. In the current report, current season influenza vaccination status was unknown for 10% of pediatric deaths compared with 19% of pediatric deaths reported during the 2010–2014 seasons.

This analysis is subject to several limitations. Cases included in this analysis may not represent all influenza related deaths. Some deaths due to influenza may not have had laboratory confirmation or may not have been reported to state health departments. Secondly, vaccination may not have been recorded in immunization records available to state and local health department personnel who completed case report forms. Misclassification of current season influenza vaccination status among vaccinated children would have overestimated vaccine effectiveness. However, influenza vaccination data and medical records were available from state immunization information systems for the majority of cases, including date of vaccination and vaccine type. Further, state surveillance personnel cross-referenced influenza vaccination data with those of routine childhood immunizations to verify record linkage. Third, underlying medical conditions may not have been documented in health records for some children and adolescents. Surveillance officers may not have received complete case medical histories, especially for children who died before hospitalization. Fourth, NHIS survey questions to estimate coverage among children with specified health conditions were limited to asthma episodes and diagnosed diabetes, and the ratio of coverage among children with versus without asthma or diabetes was assumed to be constant throughout the influenza season. If children with asthma or diabetes tend to be vaccinated earlier than other children, coverage among high-risk cohorts may have been greater in September and November, which would have underestimated effectiveness against deaths in children with underlying medical conditions.

Influenza vaccination reduced risk of influenza-associated death among children with or without underlying medical conditions. Together with recent reports on the large number of influenza-associated pediatric deaths during the 2024–2025 influenza season,^4^ and low influenza vaccination among children with influenza-associated necrotizing encephalopathy,^39-41^ findings of this analysis highlight the importance of seasonal influenza vaccination to prevent severe illness and complications of influenza in children and adolescents.

## Data Availability

All data produced in the present study are available upon reasonable request to the authors

## ACKNOWLEDGMENTS

We thank the influenza surveillance coordinators for their contributions to this study and all the clinicians, medical examiners, and local, state, and territorial health-department colleagues who contributed to the surveillance of pediatric influenza-associated deaths.

## DISCLOSURES

This work was funded by the U.S. Centers for Disease Control and Prevention. The authors have no conflicts of interest. The findings and conclusions in this report are those of the authors and do not necessarily represent the views of the U.S. Centers for Disease Control and Prevention.

## Notes

### Competing Interest Statement

The authors have declared no competing interest.

### Funding Statement

This study was funded by the U.S. Centers for Disease Control and Prevention

### Author Declarations

This activity was reviewed by the Centers for Disease Control and Prevention (CDC), deemed not research, and was conducted in accordance with applicable federal law and CDC policy.

